# Strategies and missed opportunities in achieving triple elimination of mother-to-child transmission (EMTCT) of HIV, syphilis, and hepatitis B virus: a scoping review

**DOI:** 10.1101/2025.06.26.25330370

**Authors:** Fikir Kibret, Morkor Newman Owiredu, Caitlin Quinn, Magdalena Barr-DiChiara, Olufunmilayo Lesi, Niklas Luhmann, Chelsea Morroni, Agnes Chetty, Akudo Ikpeazu, Casimir Manzengo Mingiedi, Deyer Gopinath, Cheryl C. Johnson, Aliza Monroe-Wise, Alison L. Drake

**Author notes:** Corresponding author:* Alison L. Drake.

## Abstract

Eliminating mother-to-child transmission (EMTCT) of HIV, syphilis, and hepatitis B virus (HBV) is a global public health priority. Despite progress towards reducing rates of mother-to-child transmission (MTCT) of HIV, global progress toward EMTCT of syphilis and HBV remains suboptimal. We systematically searched records in PubMed capturing the synergistic challenges and potential strategies in achieving triple EMTCT between January 1, 2013, and April 27, 2023. After screening 547 records, 50 reports met inclusion criteria. Overall, testing for HIV was well-established within antenatal care but limited for syphilis and HBV due to poor political support, inadequate funding, and insufficient operational guidance on how to deliver integrated testing within maternal and child health care. MTCT of syphilis and HBV appears to be impeded by lack of integrated systems to enable test-and-treat approaches with same-day test results and immediate treatment initiation, gaps in linkage to care, frequent stockouts of essential drugs and other commodities, prohibitive user fees, and limited clinical familiarity and training of staff in managing syphilis and HBV positive cases. To achieve triple EMTCT, co-strategies that leverage success of HIV programs to gain momentum towards prevention of congenital syphilis and MTCT of HBV, that break down siloed programs by co-delivery of testing, treatment, and training and that are supported through political will, advocacy, policy and funding are critical.

## INTRODUCTION

Triple elimination of [vertical] mother-to-child transmission (EMTCT) of HIV, syphilis, and hepatitis B virus (HBV) is a global public health priority. All three infections are key drivers of maternal and infant morbidity and mortality.^1–4^ Sequelae of perinatal infections can include immunosuppression and poor growth and development during early childhood following HIV infection, stillbirths and central nervous system abnormalities resulting from congenital syphilis, and chronic liver disease resulting from neonatal HBV. Substantial progress has been made towards EMTCT of HIV, with transmission rates as low as 2-4% in some sub-Saharan African countries, the region most impacted by the HIV.^5^ As of 2024, 16 countries have been validated for elimination of HIV and/or syphilis by the World Health Organization (WHO), one for EMTCT of HIV alone, one for syphilis alone^6^, and 14 countries have achieved dual EMTCT of HIV and syphilis; but no countries have achieved elimination of congenital HBV.^6–10^ Furthermore, no sub-Saharan country has achieved EMTCT for any of these three infections, despite the majority of infections occurring in this region. However, two countries have achieved the path to elimination designation for triple elimination, Botswana in 2021^11^ and Namibia in 2023.^12^

The rationale to combine efforts to achieve elimination for HIV, syphilis, and HBV infections is strong since these infections are all sexually and vertically transmitted, may present as asymptomatic in women or may have a long latency period, and can be identified and treated through “test-and-treat” approaches during pregnancy and breastfeeding to prevent vertical transmission.^13^ With recent funding cuts impacting services globally, integration of services is more vital than ever. Prevention of congenital syphilis and HBV may leverage successful strategies used to reduce vertical HIV transmission rates, largely focused on testing and treating pregnant and postpartum women for HIV.

Siloed approaches focused on preventing vertical HIV, syphilis, or HBV transmission independently fail to harness the potential of synergistic prevention efforts and are more costly. Identifying strategies that leverage existing systems, infrastructure, and resources—which largely exist for HIV—to achieve cross-cutting goals for the triple elimination of vertical transmission of HIV, hepatitis B, and syphilis can be cost-effective and will be critical for its success. Yet, recognizing that unique aspects of the prevention, diagnosis, and management of each infection also add complexity. Novel and innovative strategies will also be necessary to achieve triple elimination. We conducted a scoping review to gather ideas and perspectives about challenges and strategies for triple elimination.

## METHODS

### Search strategy, selection process, and data extraction

We reviewed records published in PubMed between January 1, 2013, and April 27, 2023, to identify strategies that are, or could be, used to achieve dual or triple EMTCT, as well as identify challenges and gaps. We used the following combinations of search terms: ‘(eliminate OR prevent OR testing OR diagnose OR immunization OR treat) AND (vertical transmission OR mother-to-child transmission OR infectious pregnancy complication) AND HIV AND (syphilis OR hepatitis B)’ and applied filters to restrict publications between 2013 and 2023. Additionally, we included one guidance document and one publication recently released that the authors were previously aware of which presented strategies toward triple elimination. This approach was designed not to holistically identify all prevention strategies individually for HIV, syphilis, and HBV, but rather publications that were focused on strategies and challenges for dual and triple elimination, specifically. Publications could include commentaries, editorials, reviews, briefs, guidance documents, or original research.

One reviewer (FK) screened titles and abstracts from selected titles on elimination of HIV syphilis, and/or HBV. Records were eligible for full-text review if they included information on strategies or challenges with EMTCT of HIV and at least one other infection (syphilis or HBV) and had titles and abstracts in English. Publications without abstracts (commentaries, research letters, editorials, correspondence, and research papers) were reviewed in full-text for eligibility. Full-text records were excluded from this scoping review if they reported on 1) the prevalence of HIV, syphilis, and/or HBV in pregnant women without strategies or challenges used for preventing vertical transmission; or 2) individual-level risk factors for prevention, testing, treatment, or linkage to care for of HIV, syphilis, and/or HBV.

We used a standardized form to extract data on strategies and challenges to vertical transmission, stratified by intervention categories (primary prevention, testing, treatment, and linkage to care) and population (pregnant women, infants, and partners) (Supplementary File 1). One independent reviewer (FK) reviewed the full-text articles for inclusion and exclusion and extracted relevant data into relevant categories, and a second independent reviewer (ALD) reviewed the extracted data and categorization (Supplementary Tables 1-4).Two reviewers (ALD and AMW) conducted a content analysis of the extraction sheet, and classified findings based on themes of vertical transmission strategies and missed opportunities, and created a consolidated summary as shown in Table 1.

**Table 1:** Strategies and missed opportunities for triple elimination.

|  | STRATEGIES | MISSED OPPORTUNITIES |
| --- | --- | --- |
| **Access & demand** | -Decentralize EMTCT efforts to health centers/dispensaries<br>- Educational campaigns<br>-Sensitize community about early ANC attendance through CHW<br>-Support free ANC/maternity care<br>-Achieve high rates ANC attendance and facility delivery<br>-Increase access to HBsAg RDT, HBV DNA test, HBV birth dose, TDF prophylaxis for HBV<br>-Reduce cost of HBV DNA and HBeAg tests | -Dual RDT,HBsAg RDT, and HBV DNA tests too expensive and unavailable<br>-Late presentation to ANC<br>- Misperceptions about testing<br>- Stigma<br>- Relationship and gender dynamics<br>-Lack of mechanism for HBV birth dose for non-facility deliveries |
| **Capacity building** | -Support training for providers on EMTCT of all three infections<br>- Task shifting | -Staff not trained on all 3 infections<br>-Turnover of staff within programs |
| **Testing services, lab capacity/infra structure** | -Increase availability of dual RDT for HIV/syphilis, both in facilities and in remote areas<br>-Include syphilis testing in essential ANC/PMTCT of HIV services<br>- Develop WHO prequalified triple HIV/syphilis/HBV rapid test<br>-Develop guidelines for syphilis confirmatory testing and co-locate confirmation testing services in facilities<br>- Improve laboratory capacity and turnaround time for confirmatory testing<br>-Offer HIV/syphilis testing to partners of pregnant women (written invitations, self-test distribution, weekend appointments, male-friendly clinics, home visits, provide incentives to attend)<br>-Retest for HIV/syphilis to detect new infections and prevent reinfection<br>-Incorporate eReaders to facilitate interpretation of RDT results<br>-Incorporate testing during facility delivery | - Inconsistent availability of dual HIV/syphilis RDTs<br>- Low availability of HBsAg rapid tests<br>-Capacity lacking for laboratory confirmatory testing<br>-HBeAg tests are not WHO pre-qualified<br>-Long turn-around time to receive test results, leading to suboptimal coverage<br>-Providers perceive testing to be time consuming and complicated |
| **Advocacy, policy, funding** | -Develop national guidelines, political support, dedicated national/global funding, and communication strategy for triple elimination<br>-Increase national/global advocacy | -Relative lack of advocacy, guidance, and finances for HBV and syphilis<br>-Lack of funding for dual and RDT testing supplies/kits<br>-Programs are siloed in coordination, service delivery, and funding<br>- Poor legal frameworks for sexual health |
| **Supplies & supply chain** | -Develop cohesive supply chain/procurement process for multiple tests for each infection and avoid stockouts of test kits/supplies<br>-Supply HBV birth dose via GAVI | -Siloed programs lead to lack of integrated supply chain/procurement processes<br>-Inconsistent supplies of test kits/supplies<br>-HBV birth dose not available<br>-Complex supply needs |
| **Prevention services** | -Include TDF monotherapy in HIV-related funding<br>-Policies and guidelines for HCW administration of HBV birth dose outside of facilities | - TDF monotherapy for HBV not included in HIV programme<br>-Lack of education and value of triple EMTCT for women and providers<br>-Low uptake of HBV birth dose, particularly outside of facilities |
| **Treatment services** | -Co-location of testing and treatment services<br>-Development of guidelines/protocols for treatment<br>-HCW capacity building<br>-Strengthening supply chains | -Delayed linkage to care after diagnosis<br>-Referral networks for treatment are not robust; HCW lack familiarity with protocols for partners of women with syphilis<br>- Lack of availability of BPG and TDF on sites |
| Reporting and monitoring | -Develop integrated information systems, monitoring and evaluation, surveillance, and reporting systems for dual testing<br>-Include congenital syphilis in existing data reporting systems for HIV<br>-*Improve congenital syphilis reporting* | -Insufficient staffing to collect and collate data for triple EMT |

## RESULTS

Out of 549 unique citations identified, 50 were included in this review (Figure 1). Overall, 37 represented reports from specific geographical locations [the Americas (26%, 13/50), Africa (24%, 12/50), Asia (Western Pacific: 20%, 10/50 and South-East Asia: <1%, 2/50)], while the remaining 13 reports were either cross-regional, reviews, or expert opinions. Publications broadly focused on challenges, missed opportunities, and strategies in the following eight domains: 1) access and demand; 2) capacity building; 3) testing services, laboratory capacity, and infrastructure; 4) advocacy, policy, and funding; 5) supplies and supply chain; 6) prevention services; 7) treatment services; and 8) reporting and monitoring (Table 1).

**Figure 1:**
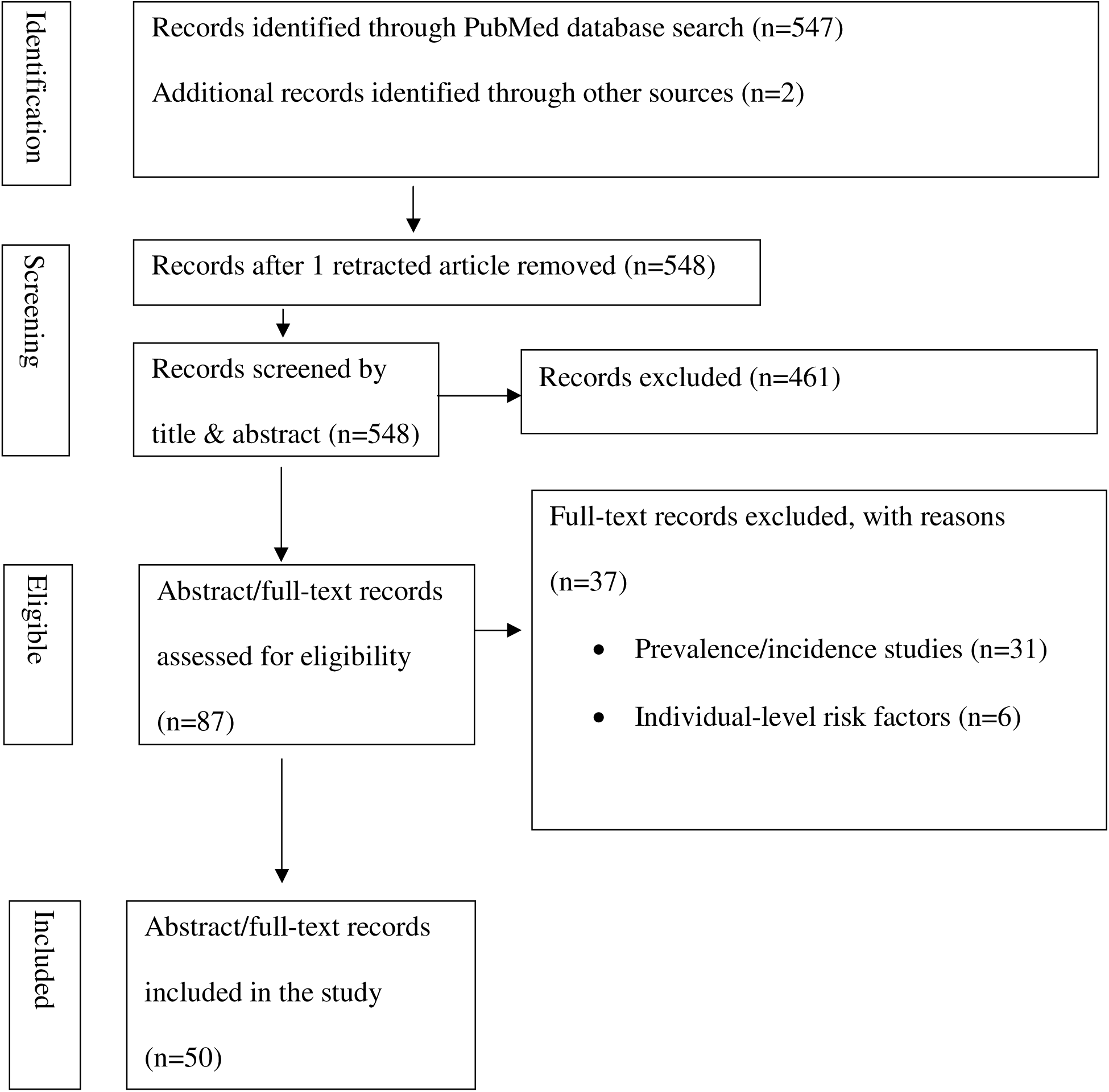
Flow diagram for record selection.

### Access and demand

#### Challenges & missed opportunities

While timely presentation to care promotes early detection, treatment, and initiation of interventions to maximize efforts to prevent MTCT of HIV, syphilis and HBV, delays in care occur for several reasons. Late first ANC attendance is one challenge.^14^ In addition to limited access to ANC services, lack of awareness among pregnant women about the risks associated with MTCT of HIV, syphilis, and HBV infections during pregnancy; the perception of testing as costly, complex, and time-consuming;^15^ and fear of encountering stigma upon receiving positive HIV or syphilis results^16^ also pose substantial barriers to testing acceptance at ANC visits. Anxiety about receiving positive test results, particularly for HIV, due to concerns about stigma, societal prejudice, or abandonment by male partners was also identified as a critical barrier to accept HIV/syphilis testing.^14,16–18^ Other aspects of partnerships that hinder testing coverage included the need for women to seek partner or spousal approval before HIV/syphilis testing in cultures considered patriarchal, and financial dependence on male partner.^16^

For HBV, challenges reported in sub-Saharan Africa extend beyond awareness into lack of testing for HBeAg and HBV DNA tests.^19^ Tests are largely too expensive and unavailable. HBV birth dose may be available in some settings, but similar to testing for HBV, lack of familiarity of HBV is compounded by the lack of availability of HBV birth doses at facilities where women deliver, as well as no clear distribution plan for women who deliver outside healthcare facilities.^19^

#### Strategies

Targeted educational campaigns to raise community awareness about the need and health benefits of early ANC visits; risks of MTCT of HIV, HBV, and syphilis; and available services to prevent MTCT of these infections have been proposed as a potential strategy that could improve rates of early ANC testing.^20^ Improving ANC access and affordability at community- and primary-care levels of healthcare systems and promoting rapid diagnostic tests (RDTs) as a strategy to facilitate same-day testing and support task-shifting might help prevent delays in seeking ANC due long distance to healthcare facilities.^20^

### Capacity building

#### Challenges & missed opportunities

Understaffing, high staff turnover, lack of familiarity of HCWs about triple elimination, and unwelcoming attitudes of healthcare providers toward pregnant women (often as a result of heavy patient loads) are also barriers to achieving EMTCT targets.^16,18–24^ Training for PMTCT of HIV has been complemented with implementation of programs that have integrated HIV testing and treatment into routine MCH services; however, providers lack knowledge and familiarity with clinical algorithms for testing and treating maternal syphilis and HBV.

#### Strategies

Broadening service provisions through task-shifting^20^ and refresher courses for HCWs^14^ have been proposed as a mechanism to improve efforts at dual HIV/syphilis elimination; however, adding content related to HBV could also help prevention efforts focus on triple elimination.^25^ Other proposed strategies to augment capacity to deliver triple EMTCT include improving equitable access to point of care testing for pregnant women (particularly in remote settings) through community healthcare workers and task shifting;^26^ increasing the proportion of deliveries with a skilled birth attendant;^21,25^ and making EMTCT services readily available to all levels of healthcare facilities, including health centers.^20^

### Testing services, laboratory capacity, and infrastructure

#### Challenges & missed opportunities

Several challenges to achieving high test coverage for HIV, syphilis, and HBV in ANC settings have been identified. The lack of availability of dual HIV/syphilis RDTs has been identified as a common challenge to achieving high syphilis test coverage, and HBsAg RDTs to detect maternal HBV infections are not widely available, making uptake at the country-level low.^19^

Additional testing is recommended for both HBV and syphilis infection after initial positive RDT results (DNA viral load or HBV envelope antigen (HBeAg) testing for HBV, and treponemal (FTA, TP-PA, EIA) testing for syphilis), and centralized laboratories are often used for these tests. However, the cost and laboratory infrastructure required to conduct these assays are significant barriers to testing; therefore, routine implementation is uncommon. ^15^ ^19^ Furthermore, the time required for confirmatory testing can be an additional barrier to linkage to care and treatment. Additionally, the absence of a WHO-prequalified HBeAg RDT product and lack of access to HBV DNA testing may contribute to low uptake of additional testing prior to antiviral prophylaxis.^19^ For both HBV and syphilis testing, out-of-pocket expenses are additional barriers to testing in some settings.^19^

For HIV-exposed infants (HEI), early infant diagnosis (EID) programs aimed at testing infants at 4-6 weeks of age can aid in identification and rapid initiation of life-saving treatment for infants who acquire HIV. Yet, loss to follow-up, inadequate health systems, testing delays, and poor social support may contribute to low EID rates among HEI, particularly in African settings where HIV prevalence is higher. ^19^ ^14^

#### Strategies

The dual HIV/syphilis RDT has been proposed as one strategy to overcome limitations with laboratory capacity and infrastructure, healthcare provider burden to administer separate tests, and to close the gap in syphilis test coverage by leveraging HIV testing infrastructure. The dual RDT has excellent clinical performance,^21,27^ is cost-effective, saves providers and patients time,^28–31^ and results in higher acceptability and test coverage compared to separate tests for HIV and syphilis.^32^ It has been recommended as a first-line HIV test in ANC by WHO since 2019 and is increasingly available in many contexts and settings. Funding for the dual RDT is covered under programs that support HIV services, which reduces barriers to financing syphilis testing in many countries,.^19^

Incorporating testing for all three infections into existing MCH programs, ideally with a single RDT, could improve antenatal testing coverage for syphilis and HBV.^33–37^ Triple HIV/syphilis/HBV RDTs are available and could be a significant intervention to improve coverage, but the quality of existing tests varies and the absence of a WHO prequalified test creates procurement challenges for countries, which limits accessibility.

To improve coverage of early EID for HIV, specialized postnatal clinics, diagnostic tests with faster turnaround times, active follow-up through advocates or health workers, and integration of testing into routine child health programs have all been proposed.^14^

Many HCWs operating within HIV PMTCT programs are aware of the benefits of referring male partners of pregnant women for HIV testing, either as individuals or couples, and treating partners who test positive for HIV. However, HCWs are less familiar with male partner testing and treatment for syphilis, making these practices more uncommon.^38^ Refresher courses for HCWs have been proposed to raise awareness about the role of partner testing and treatment for congenital syphilis prevention.^14^ Switching from an HIV RDT to a dual RDT to test male partners for both HIV and syphilis could be a strategy to build upon partner testing services for HIV and gain test coverage for partners. While some LMICs have free and universal access to syphilis treatment, including treatment of partners as an integral component of triple EMTCT,^35^ out-of-pocket fees for syphilis treatment remain as a barrier to treating partners.

### Advocacy, policy, and funding

#### Challenges & missed opportunities

Strong levels of advocacy, normative guidance, and financial support for HIV EMTCT interventions have significantly helped countries advance towards achieving EMTCT for HIV. In contrast, financial and political support for preventing congenital syphilis has been relatively lower,^16,18,21a^ and the national and global efforts to support EMTCT of HBV are substantially weaker.^19^ Specific reasons for lack of funding for syphilis and HBV EMTCT include competing health priorities, financial diversion for public health emergencies such as the COVID-19 pandemic,^39,40^ and siloed funding.^41,19,20,42^

Despite the promise of enhanced support and policies for prevention of congenital syphilis, implementation is still comparatively weak compared to PMTCT of HIV.^19,20,42,43^ Sub-optimal advocacy for congenital syphilis at national and global levels^21,24,44^ also pose threats to achieving EMTCT. In addition, triple EMTCT efforts can also be hindered by punitive laws that limit access to sexual and reproductive healthcare services for certain populations (i.e., youth and immigrants).^40^ Therefore, avoidance of coercion and legal restrictions on EMTCT service utilization may improve EMTCT efforts.^25^

#### Strategies

Political commitment and financial support are pillars to achieve EMTCT targets. For example, Cuba’s strong political commitment and dedicated national funding have been cited as enabling factors in achieving EMTCT targets for HIV and syphilis.^25^ In addition, in randomly selected health facilities that introduced an “Early Essential Newborn Care in eight countries in the Western Pacific Region, national and concerted efforts to address all three infections by leveraging existing MNCH care and integrated care, offering free testing and treatment/prophylaxis for HIV, syphilis, and HBV resulted in in HBV birth-dose coverage exceeding 80% in all but one country.^47^ However, antenatal HIV test coverage remained low (<40%) for six countries.^47^ China, however, was able to achieve 100% coverage for both maternal HIV and syphilis testing in ANC, as well as 100% coverage for HBV birth-dose and found MTCT of HIV fell over four years from 8.1% to 6.7% with strong governmental leadership and support.^33,47^

The adoption of national programs and policies, which recommend the integration of HIV/STI services within the package of ANC services,^45^ along with the commitment of several countries to adopt strategies and guidelines for eliminating congenital syphilis, has allowed countries to substantially increase syphilis testing without adversely impacting HIV testing among pregnant women in primary care facilities, and address procurement issues necessary for massive programmatic scale-up.^21,46^ Similar strategies could be employed to increase policy adoption and political support for HBV EMTCT.

### Supplies and supply chain

#### Challenges & missed opportunities

Overarching challenges affecting testing, treatment, and prevention for all three infections limit capacity to achieve EMTCT. These include siloed programs^14,24,41^ that are less efficient and lack the ability to capitalize on shared resources and prevention strategies, limited laboratory capacity, and challenges with managing separate supply chains for separate EMTCT programs. Complex supply chain management is also a challenge with the dual RDT as algorithms are required to identify women who would be ineligible to test with the dual RDT, including WLWH and/or women with a recent syphilis diagnosis, creating difficulties with forecasting for the quantity of supplies required for each test.^41^

#### Strategies

Programs could benefit from streamlining service delivery through integrated testing and treatment, overcoming problematic silos that add time, complexity, and human resource demands.^33,34,47^ Supply chains can be easier to monitor and track if they are cohesive and channeled through a universal triple elimination program, which would include factoring in complex procurement needs to test for all infections in a triple RDT, a dual test for HIV/syphilis, and multiple tests for each infection tests based on eligibility for triple or dual RDTs. This approach would help monitor inventory more closely though one program and avoid stockouts of test kits and supplies. For HBV, support from GAVI to supply the HBV birth dose is the key strategy to make meaningful inroads towards expanding access in settings where it is not available^19^.

### Prevention services

#### Challenges & missed opportunities

Oral tenofovir (TDF) is not only an effective HBV treatment but can also be used for maternal prophylaxis to prevent PMTCT. Limited availability of TDF monotherapy in LMICs has previously been shown to be problematic for HBV treatment, but is also a challenge for expanding coverage of for HB PMTCT.^19^ Furthermore, HIV programs fail to fund TDF monotherapy further hindering availability.^19^

High coverage of HBV birth dose vaccination contributes to elimination of vertical HBV transmission, but coverage remains very low in most LMICs.^19^ In a survey conducted in 16 high-burden countries in sub-Saharan Africa, only six reported providing the HBV birth dose vaccine within 24 hours of birth.^19^ HBV birth dose vaccinations are even more challenging for infants born outside of health care facilities. Few countries have policies supporting HBV birth dose vaccinations, and policy changes are necessary to incorporate HBV birth dose vaccination into national guidelines.^43^ Furthermore, Although GAVI has endorsed introducing the HBV birth dose vaccine, implementation delays and lack of availability of supplies necessary to offer these vaccines underscore the importance of improving coordination between international entities and governments to ensure a prompt and efficient rollout; and GAVI will only cover costs of implementation - not commodities.^19^ Additionally, confusion between HBV birth dose and the HBV vaccine provided as part of a 3-dose pentavalent or hexavalent vaccine series available to infants at 4-6 weeks of age have also created challenges to HBV birth dose coverage.^19,43^

Notably, our search did not return results for challenges or missed opportunities for maternal PrEP, likely due to the nature of the search which only included challenges and strategies for dual infections—HIV and syphilis, or HIV and HBV.

#### Strategies

Oral TDF is already included in first-line ART regimens for HIV treatment, and in some formulations of oral PrEP for HIV prevention (typically co-formulated with emtricitabine (TDF/FTC)), but is not included in long-acting PrEP, nor second-line ART regimens used when first-line regimens fail, or when HIV drug resistance is detected.^19^ HIV PrEP formulations that include oral TDF are commonly utilized for treating HBV infections even among HIV-negative women as it is more widely available.^19^

Policies supporting community health workers to administer the vaccine during home births could help increase HBV birth dose coverage, particularly in settings where facility delivery rates are low.^19^ Co-administration of the HBV birth dose with other birth dose vaccines, such as coupling BCG and OPV with the HBV birth dose as an integrated vaccination campaigns, has the potential to improve HBV birth dose vaccination coverage.^19^ However, while the timing of administration is at birth the mode of administration differs for all three of these vaccines (oral, intradermal, and intramuscular injection), which require different service delivery and HCW training requirements, which could present additional challenges to co-administration. Allocation of specific funding within national budgets to HBV birth dose vaccination would also augment coverage, as acknowledgement of HBV birth dose vaccination within national priorities can make vaccination very inexpensive.^19^ Finally, providing hepatitis B immunoglobulin (HBIG) to infants born to HBsAg positive mothers with high viremia is an alternative intervention available to protect infants against HBV infection, and has been shown in a modeling study to significantly lower transmission.^34^ However, HBIG is expensive, requires adequate cold chain capacity and an efficient procurement system^34^, which might limit its accessibility.

Although our review did not include results for maternal HIV PrEP implementation, many opportunities for integration exist both for HIV PrEP implementation within ANC settings, as well as within other prevention or harm reduction services.

### Treatment services

#### Challenges & missed opportunities

Delayed linkage to care following diagnosis, often due to needs for confirmatory testing, presents a significant barrier to early treatment for all three infections. Reasons for delays in linkages can also be attributed to siloed delivery of healthcare services, personnel, funding, data reporting systems, and program funding.^15^ Insufficient numbers of trained staff,^15^ as well as lack of familiarity with care and treatment of pregnant women infected with HBV and/or syphilis also contribute to these delays.^23,48^

Despite high ART coverage in many communities affected by HIV, there is limited access and uptake to benzathine penicillin G treatment for syphilis, and TDF for women with HBV who require prophylaxis or treatment.^19^ While benzathine penicillin G is inexpensive, stockouts hinder prompt treatment.^19^ Similarly, TDF monotherapy for HBV treatment is also limited in many LMICs, but is substantially more costly.^19^

#### Strategies

To overcome challenges in prompt treatment initiation and linkage to care, potential strategies may include consolidating testing and treatment services for all three infections at a common point of service delivery, financial and human resource mobilization, and providing staff training in the management of pregnant women with these infections.^20^

### Reporting and monitoring

#### Challenges & missed opportunities

Finally, strong systems for monitoring and evaluating appropriate indicators that span the triple EMTCT continuum for women, infants, and partners is imperative to measure progress and barriers to success.^35,43^ Programs with overstretched HCWs, or lack of funding to support dedicated staff for reporting, may hinder data collection and monitoring, as time and human resources to enter and monitor data may be insufficient with existing resources.

#### Strategies

Programs can invest in developing integrated information systems, supporting monitoring and evaluation activities, including surveillance as well as specific reporting systems to capture key indicators to measure progress and impact of incorporating dual testing. One strategy to improve congenital syphilis reporting that utilizes existing infrastructure without the need to develop novel systems, and improve monitoring and evaluation efforts, is the inclusion of congenital syphilis in the Global AIDS Response Progress Reporting System; this strategy could also be effective for HBV.

## DISCUSSION

Progress towards achieving triple EMTCT is inconsistent across HIV, syphilis, and HBV, with the most substantial success in reducing vertical HIV transmission. High coverage of HIV testing during routine ANC and subsequent treatment and prevention interventions have led to marked reductions in new pediatric HIV infections.^32,34–36,49^ However, gaps in testing and treatment for syphilis persist, lagging behind HIV, making dual elimination of HIV and syphilis elusive. Efforts to eliminate vertical HBV transmission also remain even more difficult, as HBV is frequently excluded in national guidelines, with subsequent low coverage of testing and treatment of pregnant women and uptake of the hepatitis B vaccine birth dose. Global reductions in funding threaten progress for elimination of vertical transmission of both syphilis and hepatitis B, and opportunities to employ cost-effective integration of services are more important than ever.

Despite evidence that integrated EMTCT services can be effective for the prevention of congenital infections, programs tasked with EMTCT of HIV, syphilis, and HBV are often fragmented and not optimized for joint planning and service integration across healthcare programs, leading to inefficiencies and higher costs. Funding disparities exist, with HIV receiving substantially more financial support than syphilis and HBV programs. Without programmatic funding to support services, expenses for syphilis and HBV EMTCT interventions are passed on to women and their families, hindering programmatic integration of care for all three infections. Political will and technical support to prioritize MTCT of HIV led to the expansion of PMTCT programs, while integration of syphilis and HBV interventions into routine ANC services and guidelines have lagged behind. Providing integrated EMTCT services for all three infections has the potential to not only help ensure streamlining of supply chains, but may also be cost-effective and efficient to implement—characteristics that are pivotal in the current funding climate.^34^

Future efforts to accelerate progress towards EMTCT of syphilis and HBV may find momentum by mirroring and/or leveraging successes from EMTCT of HIV. Community awareness, and prioritization by national governments and health care providers are necessary components of elimination efforts and help catalyze testing early in ANC. Achieving high levels of test coverage is critical to maximize subsequent elements of the treatment and prevention cascade, as demonstrated by the testing and treatment metrics used to validate EMTCT.^50^ Aggressive programs to combat HIV in areas where the epidemic was generalized have been successful, including Thailand, the first country to be validated for EMTCT by WHO in Asia,^51^ and Botswana, the first high HIV burden country to both test and treat >90% of pregnant women.^52^ Developing similar strategies as used for HIV to raise awareness, and support for testing, treatment, and prevention of congenital syphilis and HBV infection, particularly if coupled with HIV EMTCT services, has potential to accelerate triple elimination progress. Recent reductions in global public health funding, particularly for HIV and other infectious diseases, will present an additional, and significant, hurdle for countries to make strides in activities towards triple elimination. In the face of these new challenges, countries will need to develop creative financing models to simultaneously continue to support HIV prevention and treatment programs, win addition to financial support for prevention of congenital syphilis and HBV infection. De-siloed systems under universal health care and co-delivery of services could be one cost-reducing approach to move closer to these goals. However, while funding for prevention is necessary, it is not sufficient, as policies supporting procurement of commodities and resources to effectively implement programs is also essential. Investments in infrastructure for monitoring and evaluation systems to track progress towards EMTCT have previously been cited as contributing factors to achieving EMTCT of HIV,^51^ but systems will require capture and assessment of indicators specific for triple EMCT as lack of indicators have been shown to be barriers to EMTCT of HIV.^53^

Strong policies and programs supporting integration, or co-delivery of services, in ANC was one of the most influential factors contributing to dual EMTCT in Cuba, Thailand, and Belarus.^50^ Breaking down programmatic siloes to coordinate and streamline services is efficient, and momentum from one program, such as PMTCT of HIV, can help improve efforts for other integrated programs. The dual RDT has enormous potential to improve syphilis test coverage by using a single point-of-care test. However, availability remains limited, which hinders widespread use. In addition, while WHO universally recommends the dual RDT as a first line test during pregnancy, women living with HIV and those with a prior history of syphilis infection are not advised to test with the dual RDT. Therefore, providers are required to assess eligibility for the dual test and procurement plans need to address an adequate supply of multiple types of tests to meet the diverse testing needs of women, and providers need training to gain competencies in determining which test(s) to use for diagnosis for specific populations. Provider training would also assist with improving comfort and familiarity with managing syphilis and HBV infections. The addition of a triple test would likely require similar challenges with multiple procurement streams based on test eligibility but would be expected to make significant gains in HBV test coverage and overall gains towards implementation of elimination efforts.

Although strategies addressing HBV family and partner testing strategies were not found in this scoping review, these would also improve case-finding for HBV and could be integrated with family and household testing for HIV and syphilis. Furthermore, testing coverage for HBV in ANC lags behind both HIV and syphilis in many areas (however in parts of Asia HBV testing surpasses syphilis testing)^15^; however, we did not find any reports in our search addressing specific issues for HIV and HBV testing. However, we did find support for additional training on HBV for HCW to improve testing.

In addition to training for HBV testing and treatment, improvements in diagnostics and access to maternal and infant HBV prevention would be strategic investments to promote uptake. While point-of-care WHO-prequalified HBsAg RDTs exist, availability within ANC programmes is even more limited than the dual RDT. Triple RDTs are available, but they are not WHO-prequalified, and countries may only be supported in purchasing WHO pre-qualified tests under funding agreements or may lack confidence in the quality of these tests as they have not been rigorously evaluated and tested according to WHO standards. Bundling HBsAg RDTs with dual RDTs, or prequalifying multiplex RDTs that include HBsAg, has the potential to sustain progress for HIV, continue the momentum for syphilis, and catalyze prevention efforts for HBV. Advancements in rapid diagnostics that replicate desirable qualities of high quality RDTs for new triple RDT, potentially including construction of a target product profile to maximize desirable attributes across all three infections, would be expected to have high utility to advance the triple elimination agenda.

Expanding funding to holistically view and support triple EMTCT as a global health priority is worth the investment and it would be beneficial to invest in prevention interventions, such as TDF monotherapy for maternal HBV prophylaxis, BPG for maternal treatment and congenital syphilis prevention, and HBV birth dose vaccination. Although PEPFAR and The Global Fund have indicated openness to funding treatment of HBV-HIV co-infected patients, funding dedicated to TDF prophylaxis in antenatal settings is minimal. Adopting WHO recommendations to include TDF in the essential medicines list for HBV treatment within national guidelines could significantly enhance prophylactic and therapeutic options. Furthermore, considering adding TDF into long-acting PrEP formulations holds potential to provide dual protection against both HIV and HBV infection. Similarly, the addition of TDF to second-line ART regimens could prevent HBV acquisition among pregnant women living with HIV As stockouts of BPG remain a significant barrier, funding to maintain and monitor supply chains could improve access to this important prevention intervention. For infants, national immunization campaigns and policies recommending HBV birth dose administration concurrently with Bacillus Calmette-Guérin (BCG) and/or oral polio vaccines at birth could significantly bolster HBV birth dose coverage in LMICs. Finally, inclusive policies that reduce barriers to treatment initiation, including recent WHO guidelines on initiating treatment for syphilis and HBV prior to additional testing, or if additional testing is not available, can effectively reduce barriers to EMTCT.^54,55^

Many overarching challenges across the care cascade have impeded progress towards achieving triple EMTCT targets by 2030. Countries that have been validated for dual elimination of HIV and syphilis have common qualities, including equitable and universal access to healthcare, strong political commitment and increased domestic funding for programs, and efficient integrated service delivery models. Synchronizing EMTCT interventions for all three infections within maternal and child health (MCH) platforms has high potential to enhance efficiencies in providing comprehensive EMTCT services, which is key to propel countries closer to achieving triple EMTCT.

Our review has many strengths. It provides a contemporary view of the current strategies and missed opportunities towards triple EMTCT in LMIC settings. We included scientific research as well as commentaries and editorials with perspectives and recommendations of where future efforts could be directed to achieve triple EMTCT. In addition, our review focused on articles examining elimination of at least two infections rather than PMTCT of individual infections to examine synergistic strategies and missed opportunities across infections.

Our review is also subject to some limitations. We restricted our search based on language, geographic location, and only included records from the last ten years, which may bias our results or omit relevant findings. In addition, several records included did not clearly define “integration,” and limited our ability to clearly summarize findings and recommendations. Furthermore, we did not review the grey literature where reports, guidance documents, or programmatic findings may have been presented. In addition, there were insufficient numbers of articles to adequately to compare and contrast findings across different contexts and settings, and findings are presented with limited reference to geographic setting despite the potential for there to be key differences in different settings. Finally, we may have missed strategies or challenges towards EMTCT of individual infections by excluding articles that were limited to only one infection. For example, some highly effective prevention interventions, such as HIV PrEP were notably absent in our search as most of the HIV PrEP literature is focused on HIV alone, rather than dual or triple EMTCT.

In conclusion, implementation and scale-up of several interventions aimed at preventing MTCT of HIV have shown remarkable progress in global efforts towards elimination of MTCT of HIV: yet, significant challenges remain for EMTCT of syphilis and HBV within the changing global health landscape. Success and progress in the EMTCT cascade for HIV for prevention, testing, treatment, and linkage to care should be leveraged to include similar strategies for syphilis and HBV. International and national organizations should intensify their political, funding, and technical support to accelerating progress towards global triple EMTCT goals, including investments in technologies such as a triple RDT that can simultaneously test for all three infections.

## Data Availability

All data produced in the present study are available upon reasonable request to the authors

## Authors’ contributions

FK developed and implemented the search. FK and ALD extracted data. FK, ALD, and CQ designed the protocol. FK, AMW, and ALD synthesized the data. FK, ALD, AMW wrote the manuscript. FK, ALD, MNO, CQ, MB, OL, NL, CM, PMAS, AC, AI, EO, CMM, DG, KL, CCJ, and AMW all provided substantial input and edits to the manuscript.

## Acknowledgments

We acknowledge support from the University of Washington’s Global Center for Integrated Health of Women, Adolescents, and Children (Global WACh). The findings and conclusions in this report are those of the authors and do not necessarily represent the official position of the World Health Organization.

---

**Table 4:**
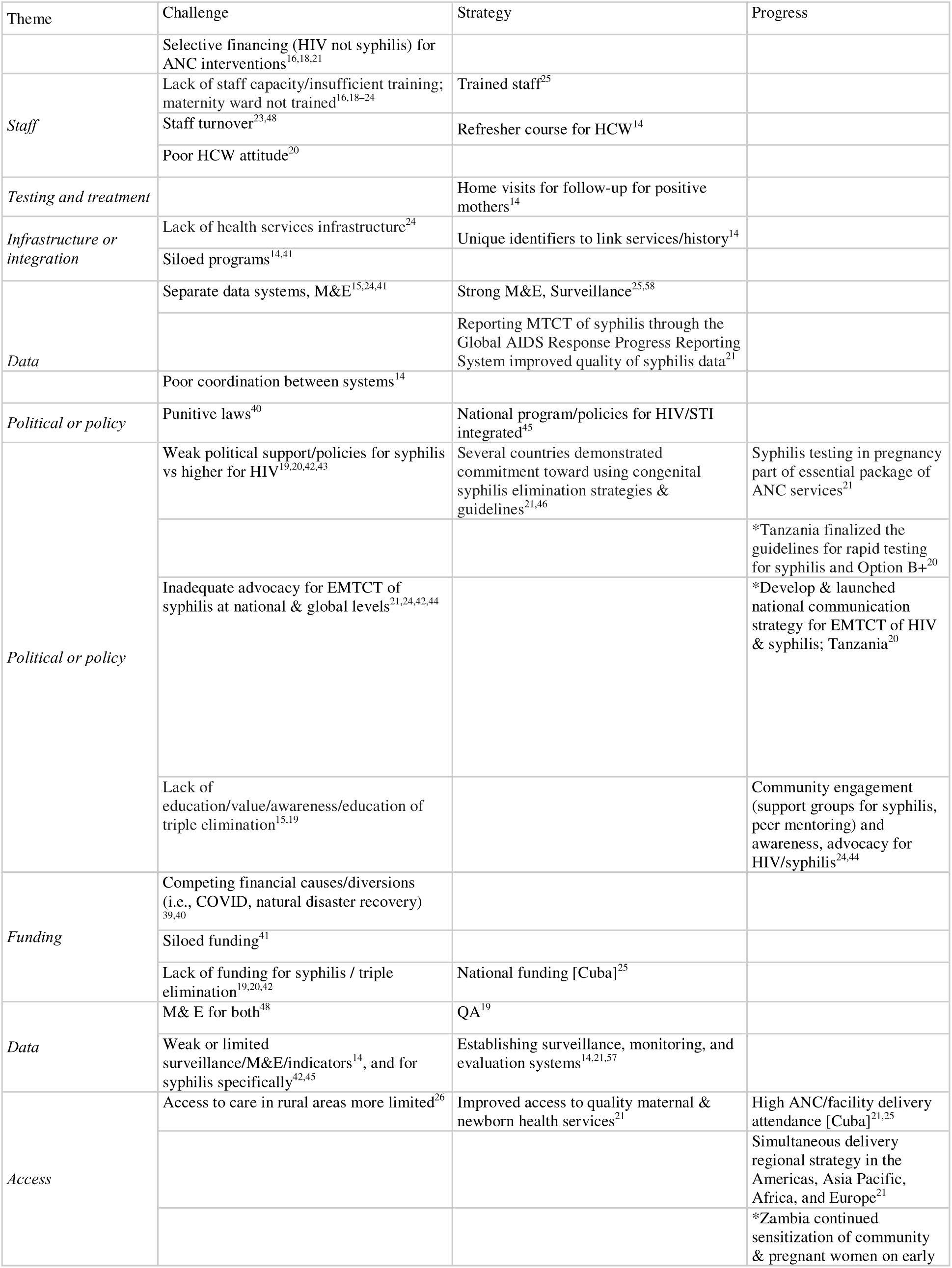
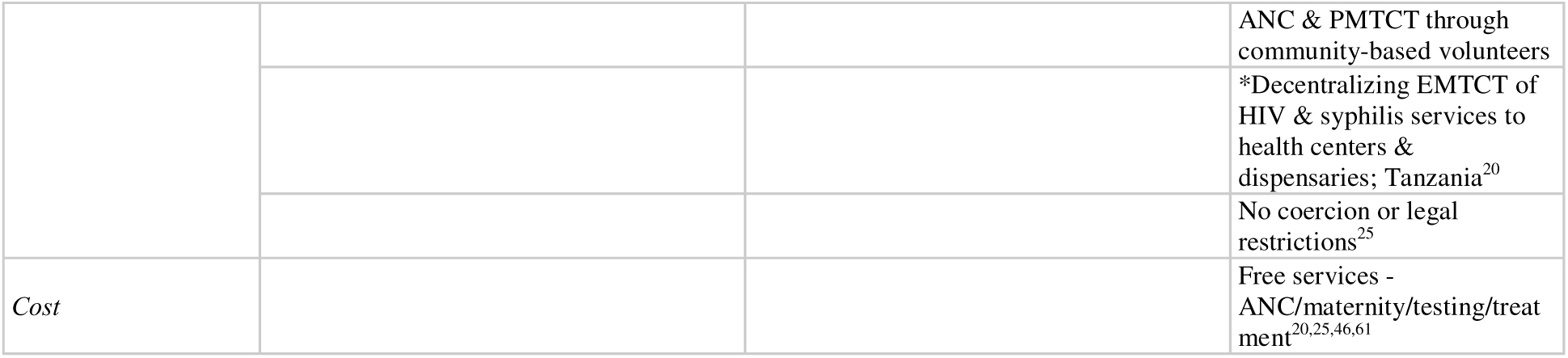
General challenges, strategies, and progress in EMTCT.

**Supplementary Table 1:** Challenges, strategies, and progresses in testing to achieve triple EMTCT.

| Theme | Challenge | Strategy | Progress |
| --- | --- | --- | --- |
| *Dual test* | Dual HIV/Syphilis RDT suboptimal availability<sup>19</sup> | Dual HIV/Syphilis RDT<sup>19,20</sup> | Dual test increases test coverage<sup>32</sup> |
|  |  | Dual RDT excellent clinical performance<sup>21,27</sup> | Dual RDT increased % timely treated for syphilis<sup>56</sup> |
| *Triple test* | HBsAg RDT not widely available<sup>19</sup> |  |  |
|  | HBeAg less accurate, not widely available, more complex to test, not WHO pre-qualified test<sup>19</sup> |  |  |
|  | HBV DNA follow up test not widely available & expensive<sup>19</sup> |  |  |
|  | Supply chain/procurement processes different for each infection<sup>19</sup> | WHO pre-qualified HIV/syphilis/HBsAg RDT |  |
|  |  | Triple RDT may be donor eligible (includes HIV), similar to dual |  |
| *General testing* | Delayed test results or delivery of test results<sup>14,20</sup> | Concurrent testing<sup>48</sup> | Concurrent testing increased syphilis testing in Kenya<sup>48</sup> |
|  |  | Same-day diagnosis and treatment<sup>14</sup> |  |
|  | Suboptimal testing and treatment [ India], testing 83% HIV, 46% syphilis, treatment 64% HIV, 47% syphilis; MTCT 24%<sup>20,57</sup> |  | High HIV test coverage in ANC (Xia, 2015) □ |
|  |  |  | POC testing has made syphilis testing possible even in remote settings without access to traditional labs<sup>21</sup> |
| *Testing integration* | Siloed testing<sup>15</sup> | Co-location of confirmatory testing could improve diagnosis/treatment uptake<sup>15</sup> | *Integrate syphilis testing into existing PMTCT health services; Madagascar<sup>20</sup> |
|  | Laboratory tests required for syphilis and/or lack of quality assurance<sup>19,45</sup> |  |  |
|  | Lack of laboratory preparedness<sup>24</sup> | RDT does not require lab referral<sup>18</sup> | Lab capacity/infrastructure<sup>25</sup> |
|  |  | Task shifting<sup>20</sup> | Integration improves test coverage for HIV/syphilis<sup>19</sup> |
|  | Lack of universal testing<sup>40</sup> | Universal testing of pregnant women<sup>21,58</sup> |  |
|  |  | POC smartphone test with high test performance, lower cost than ELISA, fast turnaround time. Tested in Rwanda. Highly acceptable<sup>59</sup> |  |
|  | Interpretation of RDT results delayed, QA issues (Wedderburn 2015) □ | e-Readers to interpret RDT-standardized interpretation of RDT results<sup>60</sup><br>*real-time data transmission<br>*improved quality assurance |  |
|  | Stigma for syphilis<sup>42</sup> |  |  |
| *Stigma* | Frequent RDT stockouts/test supplies for syphilis testing<sup>14,16,18,20-23</sup> |  |  |
| *Stockouts* | Mandatory testing, barrier<sup>16</sup> | Mandatory testing, strategy in Cuba [Cuba]<sup>61</sup> |  |
| *Mandates* | Lack of retesting in pregnancy, seroconversion and reinfection not detected<sup>14,22</sup> | Retesting for HIV/syphilis in pregnancy/BF<sup>22,57</sup> | HIV retesting coverage higher in higher HIV burden settings<sup>19</sup> |
|  |  | Testing at L&D (retesting)<sup>58</sup> |  |
| *Retesting* | Cost of dual HIV/syphilis RDT<sup>19,56</sup> | Dual test cost-saving or cost-effective - even if dual RDT kit is more expensive<sup>28-31</sup> | Dual RDTs cost-effective<sup>21,62</sup> |
| *Cost/time* | Testing perceived to be time consuming and complicated<sup>15</sup> |  | Dual test time saving for women and HCW, acceptable/feasible<sup>18,21,31,32,62,63</sup> |
|  | Inadequate funding for dual/RDT testing supplies/kits<sup>16,21</sup> |  | Testing for HBV cost-effective and time-saving for HCW<sup>34</sup> |
|  | Separate testing algorithms<sup>41</sup> |  |  |
| *Processes* | Lack of same day testing<sup>20</sup> |  |  |
|  | Late ANC attendance;<br>(HBV needed before 28-weeks of gestation)<sup>18,20</sup> | Early/any ANC<sup>14,20</sup> |  |
|  |  | Training<sup>48</sup> |  |
| *Staff* |  | Male partner involvement in MCH programs<br>(written invitations, weekend openings,<br>incentives, male-friendly clinics, and home visits<br>for male testing and education)<sup>44,45</sup> |  |
| *Partner* |  | Include men in ANC (PAHO 2014) ☐ |  |
| *Political*<br>*/policies* | Lack of guidelines<sup>16</sup> | At least 60 countries have integrated strategies<sup>21</sup> |  |

**Supplementary Table 2:** Challenges, strategies, and progress in treatment and linkage to care to achieve EMTCT.

|  | Theme | Challenge | Strategy | Progress |
| --- | --- | --- | --- | --- |
| MATERNAL TREATMENT AND PMTCT | *Partner* |  | Discuss potential health consequences to the baby with father/[partner] if the mother refuse treatment - Cuba<sup>61</sup> |  |
|  | *Treatment* | Lack of same day treatment<sup>20</sup> | PMTCT ART for HIV-exposed infants after birth<sup>19</sup> |  |
|  |  | Gaps in referrals for treatment<sup>15,38</sup> |  |  |
|  |  | Benzathine penicillin G stockouts for syphilis<sup>19</sup> | Benzathine penicillin G x 3 weekly for syphilis<sup>19</sup> |  |
|  |  |  | Prompt linkage to care<sup>40</sup> |  |
|  | *PMTCT* | Formulation of second line ART regimens for HIV+ do not include TDF, does not protect infants against HBV infection<sup>19</sup> |  | Treatment for HBV for women with high HBV viral load cost-effective and time-saving for HCW<sup>34</sup> |
| INFANT TREATMENT AND CARE | *Infant testing/treatment* |  | HBIG for infants born to women with HBsAg+ results within 12 hours of birth<sup>64</sup> | HBIG for HBV for infants born to HBsAg+ women cost-effective and time-saving for HCW<sup>34,64</sup> |
|  |  | Inadequate follow-up of exposed infants, low EID coverage<sup>19,38</sup> | EID and treatment if positive for HIV<sup>14,19</sup> |  |
|  |  | No policies for introducing HBV vaccine<sup>43</sup> | Include HBV birth dose vaccination in national priorities<sup>19</sup> |  |
|  | *Policy* | GAVI approved introduction of HBV birth dose, but implementation delayed; GAVI is not supplying though<sup>19</sup> |  |  |
|  |  | No dedicated funding for HBV vaccine<sup>43</sup> | Include HBV birth dose vaccination in national budget; inexpensive<sup>19</sup> |  |
|  | *Funding* | Moderate coverage for HIV, low coverage for syphilis<sup>19</sup> | Promote partner testing/couples testing<sup>14</sup> |  |
| PARTNER TREATMENT AND CARE | *Testing* | Referring partners of positive women for testing<sup>23</sup> | Offering both HIV and syphilis testing to partners was acceptable<sup>65</sup> |  |
|  |  | Lack of awareness for HCW to refer and treat partners of pregnant women with syphilis<sup>38</sup> | Presumptive (or after confirmed positive test) partner syphilis treatment, prevents reinfection, avoids needing to test<sup>19</sup> |  |
|  | *Treatment* | Stockouts of syphilis treatment<sup>23</sup> | Promote partner testing/couples testing<sup>14</sup> |  |
|  |  |  | Home based testing for partners for HIV and syphilis acceptable<sup>65</sup> |  |
|  |  | Syphilis: Benzathine penicillin G stockouts<sup>19</sup> | Universal treatment of positive partners<sup>14</sup> |  |

**Supplementary Table 3:** Challenges, strategies, and progress in prevention.

|  | Theme | Challenge | Strategy | Progress |
| --- | --- | --- | --- | --- |
| INFANT HBV VACCINATION |  | Limited implementation support<sup>19</sup> | HBV birth dose vaccination <24 hours; especially important if mother HBeAg+ or high HBV VL, but recommended for all regardless of mother's HBV status<sup>19</sup> | HBV birth dose vaccination coverage low<sup>19</sup> |
|  |  | HBV birth dose vaccination difficult if non-facility delivery<sup>19</sup> | HBV birth dose vaccination delivered by CHW/TBA if non-facility delivery<sup>19</sup> |  |
|  |  | Confusion between the HBV vaccine series and birth dose | Combine HBV birth dose vaccination couple with other birth vaccines (polio/BCG)<sup>19</sup> |  |
| PREVENTION (PrEP) | Access | TDF prophylaxis coverage for HBV low, even if in policy<sup>19</sup> |  |  |
|  |  | TDF monotherapy availability for HBV low<sup>19</sup> | TDF monotherapy or 2-drug (formulated as HIV PrEP) for HBV PrEP if high HBV VL and at high risk of acquiring HIV-; offers added protection against HIV acquisition<sup>19</sup> |  |
|  |  | Long-acting PrEP for HIV does not contain TDF, does not protect against HBV infection<sup>19</sup> | ART for women living HIV with TDF (included as first line regimen) which protects against maternal HBV infection<sup>19</sup> |  |
|  | Funding | TDF monotherapy for HBV not covered by HIV funders<sup>19</sup> |  |  |
|  | General |  | PrEP for HIV<sup>19</sup> |  |
| SRH |  |  | Add HIV/syphilis testing into FP programs (postpartum)<sup>44</sup> |  |
|  |  |  | ART for HIV prior to pregnancy<sup>19</sup> |  |
|  |  |  | Expand access to FP<sup>14</sup> |  |

